# Feeding practices and cost of diet of six-month-old HIV exposed uninfected compared to HIV unexposed uninfected infants in a peri-urban community in South Africa

**DOI:** 10.1101/2022.03.28.22273056

**Authors:** Mothusi Nyofane, Marinel Hoffman, Tanita Botha, Ute Feucht

## Abstract

**Background:** The increasing population of HIV-exposed-uninfected (HEU) infants are known to be at risk of poor nutritional status and suboptimal growth, with biological risk factors implicated, yet the cost to families of feeding infants is often overlooked.

**Objective:** The study compared the infant feeding practices and costs and macronutrient intake of HEU vs HIV-unexposed-uninfected (HUU) six-month-old infants in the Gauteng Province of South Africa.

**Methods:** A cross-sectional study investigated 46 HEU and 55 HUU infants aged six months and utilised a single quantified 24-hr recall and the FoodFinder™ program for meal analysis. The estimation of diet cost utilised supermarket food prices based on the 24-hr recall method.

**Results:** Mothers of HEU infants had significantly lower income (*p*<0.01) and educational attainment (*p*=0.03). The infant feeding practices differed between HEU vs HUU infants (*p*=0.05): exclusive breastfeeding (50.0% vs 34.0%) and mixed breastfeeding (38.1% vs 64.2%). Common complementary foods for HEU versus HUU infants included commercial infant cereals (CIC) (48.7% vs 70.9%; *p*=0.04); fruits and vegetables (33.3% vs 15.7%; *p*=0.05) and maize meal porridge (25.6% vs 15.7%; *p*=0.24), respectively. The mean daily cost of diet of HEU vs HUU infants was 8.55±7.35ZAR ($0.68±0.59USD) vs 10.97±7.92ZAR($0.88±0.63 USD); (*p*=0.10). Regarding the complementary feeding, there were non-significant differences in protein, fat, and carbohydrate intakes (*p*>0.05) and their costs per daily intake (*p*>0.05) between the groups.

**Conclusion:** There are no significant differences in cost, feeding and macronutrient intakes between HEU and HUU. Suboptimal breastfeeding practices remains an issue within the first six months. More sustained effort is required to support and promote exclusive breastfeeding.

## Introduction

Globally, the human immunodeficiency virus (HIV) continues to adversely affect women, in particular, women of reproductive age [1]. Approximately 1.4 million women living with HIV infection (WLHIV) become pregnant every year [2]. South Africa is greatly affected, with 8.2 million people living with HIV in 2021 [3], with a high HIV prevalence (26.3%) among women of reproductive age [4]. In the absence of antiretroviral therapy (ART), there is a 25% to 30% risk of mother to child transmission of HIV during pregnancy, delivery and breastfeeding [1]. The World Health Organization (WHO) prevention of mother-to-child transmission (PMTCT) guideline recommends that all pregnant WLHIV should be on ART during pregnancy as part of PMTCT interventions [1,5]. In South Africa, access to ART has increased over the years, with an estimated >95% ART coverage during pregnancy and delivery [6], and this has ensured sharp declines in new vertical HIV transmissions in infants born to WLHIV [2,6,7,8]. This has resulted in an expanding population of HIV-exposed-uninfected (HEU) infants, estimated globally at 14.8 million in 2018, with South Africa having a high HEU population of 3.5 million (23.8%) [2,9]. Compared to their HIV-unexposed-uninfected (HUU) counterparts, the HEU children’s early and later growth has been reported to be suboptimal [10], with the risk of malnutrition [11].

The 2016 WHO guideline update on HIV and infant feeding advised alignment of infant feeding guidelines for WLHIV and HIV-negative women within the context of ART provision for WLHIV [12]. These include exclusive breastfeeding (EBF) for the first six months, with subsequent introduction of high-quality complementary feeds together with continued breastfeeding [12]. Nonetheless, the percentage of EBF is still suboptimal in South Africa. The South African Demographic and Health Survey of 2016 reported EBF of 32% among children under age six months [13], and Remmert et al. reported 30% EBF among WLHIV in KwaZulu-Natal, South Africa [14]. The maternal HIV status has an influence on breastfeeding practices, including breastfeeding cessation due to fear of HIV vertical transmission [14,15]. The suboptimal breastfeeding practices, coupled with early and/or inadequate introduction of complementary feeding, is associated with a high risk of infant infections and malnutrition [16,17].

Infants require a healthy diet at the proper time to grow and develop to their full potential. There is a strong reported relationship between HIV infection, poverty, food insecurity, and malnutrition, with suboptimal maternal health resulting in reduced ability to perform daily activities, including income generation, adequate infant care, and feeding practices [18]. South African complementary foods mostly comprise of maize meal porridge and commercial infant cereals (CIC) [19,20,21]. Food prices strongly influence the selection of food and dietary quality and ultimately affect infant nutritional status [22,23]. The CIC have a high nutrient density but are more costly than low nutrient density staple foods used as complementary feeds [19,24]. Macronutrient intake plays a central role during infant growth and development and throughout life. Fatty acids support growth and development through their roles in membrane lipids and supporting brain and retinal development; thus, insufficient intake limits neurodevelopment [25]. Protein intake is vital for health, physical growth and development, with dietary protein deficiency causing different forms of undernutrition, such as stunting and severe acute malnutrition [26]. These macronutrients, together with carbohydrates, provide energy and building blocks essential for growth.

The WHO recommends the introduction of age-appropriate complementary feeds at the age of six months after initial EBF [27], but little is documented about the cost of diet to the families and the comparison in infant feeding practices between HEU and HUU infants. Better knowledge on real-life infant feeding practices can enhance guidance given to mothers on infant feeding practices, food choices and complementary diet planning. Cost of diet analysis allows identification of food items contributing to nutrient-dense diet and their cost, including food items that contribute to high cost but with low nutrients. We therefore determined and compared the cost of diet in relation to nutrient intake and feeding practices of HEU versus HUU infants aged six months.

## Materials and Methods

### Study design

This cross-sectional and comparative study was part of a larger study (Siyakhula study) on evaluating factors affecting foetal and infant immunity, growth, and neurodevelopment in HEU children. Low-risk (as per South African antenatal care guidelines) pregnant WLHIV and HIV-negative women were identified from 2017 and continually recruited at 28 weeks of pregnancy at the University of Pretoria/South African Medical Research Council (SAMRC) Research Centre for Maternal, Fetal, Newborn and Child Health Care Strategies in Kalafong Provincial Tertiary Hospital in Gauteng Province, South Africa, with their HEU or HUU infants being longitudinally followed up for two years. The infants started turning six months from October 2018 and data collection for this study was completed in May 2020.

### Study population

Participants were screened for eligibility by research assistants in local languages and English. Exclusion criteria included mothers who were minors (<18 years), had multiple pregnancies, maternal chronic medical conditions including diabetes and hypertension, HIV-infected infants, and lack of informed consent. The anticipated sample size was 100 mother-infant dyads (HEU n=50; HUU n=50). In May 2020, 113 participants had six-month-old infants; of these, 12 mother-infant dyads had either lost to follow-up to the study or relocated (n=11), or infants had been diagnosed with HIV infection (n=1). Therefore, 101 mother-infant dyads (HEU n=46; HUU n=55) were investigated. All WLHIV self-reported taking ART during pregnancy and postpartum. Infants (HEU n=7; HUU n=4) who were exclusively breastfed at the time of study visit were excluded in the cost of diet analysis. Therefore, cost of diet was calculated for 39 HEU and 51 HUU infants.

### Data collection

Written informed consent was obtained from eligible mothers for themselves and their infants to participate in the study. The researcher and trained research assistants administered the questionnaires in a private room in local languages and English, and mothers were compensated with 100 ZAR (roughly 7 USD) for transport. Interviews lasted for 30–40 minutes. The quantified single 24-hr recall was used to collect infant dietary intake and included intake on Sundays, Tuesdays, and Wednesdays. The research assistants were trained on the procedures for administering a 24-hr recall. For estimating and recording the reported amount of consumed food, a standardized dietary kit containing samples of food and food containers, household utensils, and photographs was utilized. Dry oats were used in dish-up and measure for estimating and recording the consumed quantity, mostly for prepared food. Breast milk substitutes (BMS) and CIC were noted per dry quantity and liquid. The 24-hr recall has been validated for accurate intake measurement in children, in which the mother is interviewed to report a child’s dietary intake [28].

The estimation of diet cost utilised supermarket food prices and the diet diaries method [29], however, a single 24-hr recall was used as an alternative to diet diaries. The supermarket food prices were collected in February 2019 from three local supermarkets and the list comprised food items that were not on special offer and were of the lowest prices [29]. The previously used maternal and infant postpartum questionnaire was used to collect data on breastfeeding practices, the introduction of complementary foods and socio-demographic variables. The food purchasing practices were self-reported using a structured and pretested self-designed food cost questionnaire; however, this questionnaire was administered to a portion of participants (HEU n=23; HUU n=27).

### Data processing and statistical analysis

The collected data were managed using the Research Electronic Data Capture (REDCap) system. All household food intake measurements coded on the 24-hr recall were converted to weight using the SAMRC Food Quantities Manual prior to meal analysis and the SAMRC FoodFinder™ 3.0 program was used to quantify macronutrients intake [30]. The dietary intake included food, beverages and BMS consumed by infants. Where the flavour of food, fruit eaten or type of soft porridge drank were not specified, the commonly reported consumed food flavour (banana flavour), fruit (banana) or soft porridge (maize meal porridge) were used. Three food prices per food item were manually converted to a price per 100g of consumed food items, and the average price was calculated. The costs of diet and nutrient intake were calculated using the amount of food items eaten and macronutrients to food prices per 100g. Any intake of breastmilk was excluded, as this research aimed to describe the nutrient intake from purchased food items and BMS. The study used IBM SPSS Statistics (version 25) software for descriptive statistics. Nutrient intakes and their costs were tested for normality using the Shapiro–Wilks test. These data were not normally distributed, hence reported by the median and interquartile range (IQR). To determine the significance of differences between the HEU and HUU infants, the Mann–Whitney U test was used. All significance tests were performed at a 5% level of significance. Cessation of breastfeeding is reported categorically within the six-month period, and the statistic test was done using Pearson’s Chi-squared test.

### Ethical consideration

The study was approved by the University of Pretoria Faculty of Natural and Agricultural Sciences Research Ethics Committee and Faculty of Health Sciences Research Ethics Committee. Further permissions were obtained from Kalafong Provincial Tertiary Hospital and Tshwane District Health Department. The process of data collection was voluntary and confidential. The study was conducted in accordance with ethical guidelines set in the 2013 Declaration of Helsinki.

## Results

### Socio-demographic and food purchasing information

Significantly higher percentages of WLHIV had lower educational qualifications (52.2% vs 29.1%; *p*=0.03) and were in the lowest income category (<4000 ZAR/month; <$320 USD/month) (69.8% vs 36.4%; *p*=0.002), whereas more of the HIV-negative mothers were in the highest monthly income category of >8000 ZAR/month (>$640 USD/month) (26% vs 5%; *p*=0.002) (Table 1). We found no significant differences in food purchasing practices between the two groups.

**Table 1:** The socio-demographic characteristics and food purchasing practices of mothers, grouped by HIV infection status

|  | HIV-infected<br>mothers (n=46) | HIV-uninfected<br>mothers (n=55) | P-value<sup>a</sup> |
| --- | --- | --- | --- |
| **Age of infants (months; days) (median (IQR))** | 6.3 (6.1;6.5) | 6.2 (6.1;6.4) | 0.56 |
| **Sex of infants (%)** |  |  | 0.10 |
| Male | 30 (65.2%) | 27 (49.1%) |  |
| Female | 16 (34.8%) | 28 (50.9%) |  |
| **The educational level of the mother (%)** |  |  | **0.03** |
| Any primary or secondary school | 24 (52.2%) | 16 (29.1%) |  |
| Secondary school completed | 13 (28.3%) | 22 (40.0%) |  |
| Certificate/Diploma completed | 9 (19.6%) | 12 (21.8%) |  |
| Undergraduate degree completed | 0 (0.0%) | 5 (9.1%) |  |
| **Number of children in the household<sup>b</sup> (%)** |  |  | 0.80 |
| One or two children | 14 (66.7%) | 17 (63.0%) |  |
| Three to five children | 7 (33.3%) | 10 (37.0%) |  |
| **Household income per month<sup>c</sup> (%)** |  |  | **0.002** |
| ZAR 0 – ZAR 4000<sup>d</sup> | 30 (69.8%) | 20 (36.4%) |  |
| ZAR 4000 – ZAR 8000 | 11 (25.6%) | 21 (38.1%) |  |
| >ZAR 8000 | 2 (4.6%) | 14 (25.5%) |  |
|  | HIV-infected<br>mothers (n=23)<sup>e</sup> | HIV-uninfected<br>mothers (n=27)<sup>e</sup> | P-value |
| **Number of household members with income (%)** |  |  | **0.05** |
| One | 12 (52.2%) | 15 (55.6%) |  |
| Two | 3 (13.0%) | 8 (29.6%) |  |
| >Two | 1 (4.4%) | 3 (11.1%) |  |
| None | 7 (30.4%) | 1 (3.7%) |  |
| **Household receiving a social grant(s) (%)** |  |  | 0.92 |
| Yes, child support grant | 19 (82.6%) | 22 (81.5%) |  |
| No | 4 (17.4%) | 5 (18.5%) |  |
| **Number of children receiving grants<sup>f</sup> (%)** |  |  | 0.92 |
| One to two | 12 (66.7%) | 15 (68.2%) |  |
| ≥Three | 6 (33.3%) | 7 (31.8%) |  |
| **Food purchased in bulk or daily (%)** |  |  | 0.31 |
| Bulk | 18 (78.3%) | 24 (88.9%) |  |
| Daily | 5 (21.7%) | 3 (11.1%) |  |
| **Food purchased when on special offer (%)** |  |  | 0.56 |
| Yes | 6 (26.1%) | 5 (18.5%) |  |
| No | 4 (17.4%) | 8 (29.6%) |  |
| Sometimes | 13 (56.5%) | 14 (51.9%) |  |
| **Food purchased from<sup>g</sup> (%)** |  |  | 0.64 |
| Street vendor | 1 (4.3%) | 2 (7.7%) |  |
| Big hypermarkets/supermarkets | 2 (8.7%) | 4 (15.4%) |  |
| Local shops | 20 (87.0%) | 20 (76.9%) |  |
| **Reason for shop preference (%)** |  |  | 0.12 |
| Proximity to home | 11 (47.8%) | 6 (22.2%) |  |
| Prices of foodstuff | 7 (30.4%) | 15 (55.6%) |  |
| Quality of foodstuff | 5 (21.8%) | 6 (22.2%) |  |
| **Quality of fruits and vegetables sold on streets (%)** |  |  | 0.89 |
| Is fresh most of the times | 8 (34.8%) | 7 (25.9%) |  |
| Is sometimes fresh | 13 (56.6%) | 17 (63.0%) |  |
| Is never fresh | 1 (4.3%) | 2 (7.4%) |  |
| Not sure | 1 (4.3%) | 1 (3.7%) |  |

Abbreviations: IQR: Interquartile range
<sup>a</sup>P-values <0.05 show significant differences and are bolded. Pearson's Chi-squared was used to compare HIV-infected and HIV-uninfected mothers.
<sup>b</sup>n= 21 (HIV-infected mothers) and n= 27 (HIV-uninfected mothers)
<sup>c</sup>n= 42 (HIV-infected mothers) and n= 55 (HIV-uninfected mothers)
<sup>d</sup>Conversion factor: 1 ZAR= 0,064176 USD (Available at https://www.curvert.com/en/zar-usd/ (Cited: 13/11/2020)
<sup>e</sup>The food cost questionnaire was utilized in a subgroup of participants, which included 23 HEU and 27 HUU mother-infant pairs.
<sup>f</sup>n= 18 (HIV-infected mothers) and n= 22 (HIV-uninfected mothers)
<sup>g</sup>n= 23 (HIV-infected mothers) and n= 26 (HIV-uninfected mothers)

### Infant feeding practices

The feeding practices are presented in Table 2. The overall infant feeding practices within the first six months differed between WLHIV and their HIV-negative counterparts (*p*=0.05); the question asked, “*Thinking about the time between when your baby was born and now (this visit), how did you feed your baby from birth until now?”*. Any current breastfeeding was documented in 61.5% of WLHIV and 67.9% of HIV-negative mothers (*p*=0.37). In a sizable percentage, both WLHIV and HIV-negative women had stopped breastfeeding very early (before four weeks) (26.7% vs 11.8%; *p*=0.39). Many WLHIV started introducing BMS from as early as 4 – 8 weeks (31.3%), while HIV-negative women started giving BMS from 8 – 12 weeks (35.3%); *p*=0.49. Reasons given for the early introduction of BMS in WLHIV and their HIV-negative counterparts was because they wanted to return to work (25.0% vs 37.4%), job searching and schooling (25.0% vs 27.3%) and perceived insufficient breastmilk (25.0% vs 18.2%) (*p*=0.38). Further, the percentages of EBF during the first six months were 50.0% among WLHIV and 34.0% among HIV-negative mothers (*p*=0.20). The mixed breastfeeding (MBF) practices were higher among HIV-negative mothers (64.2%) compared to WLHIV (38.1%); (*p*=0.03). The percentages of formula feeding (FF) as only milk feeds given within the first six months were 11.9% in WLHIV and 1.8% among HIV-negative mothers.

**Table 2:**
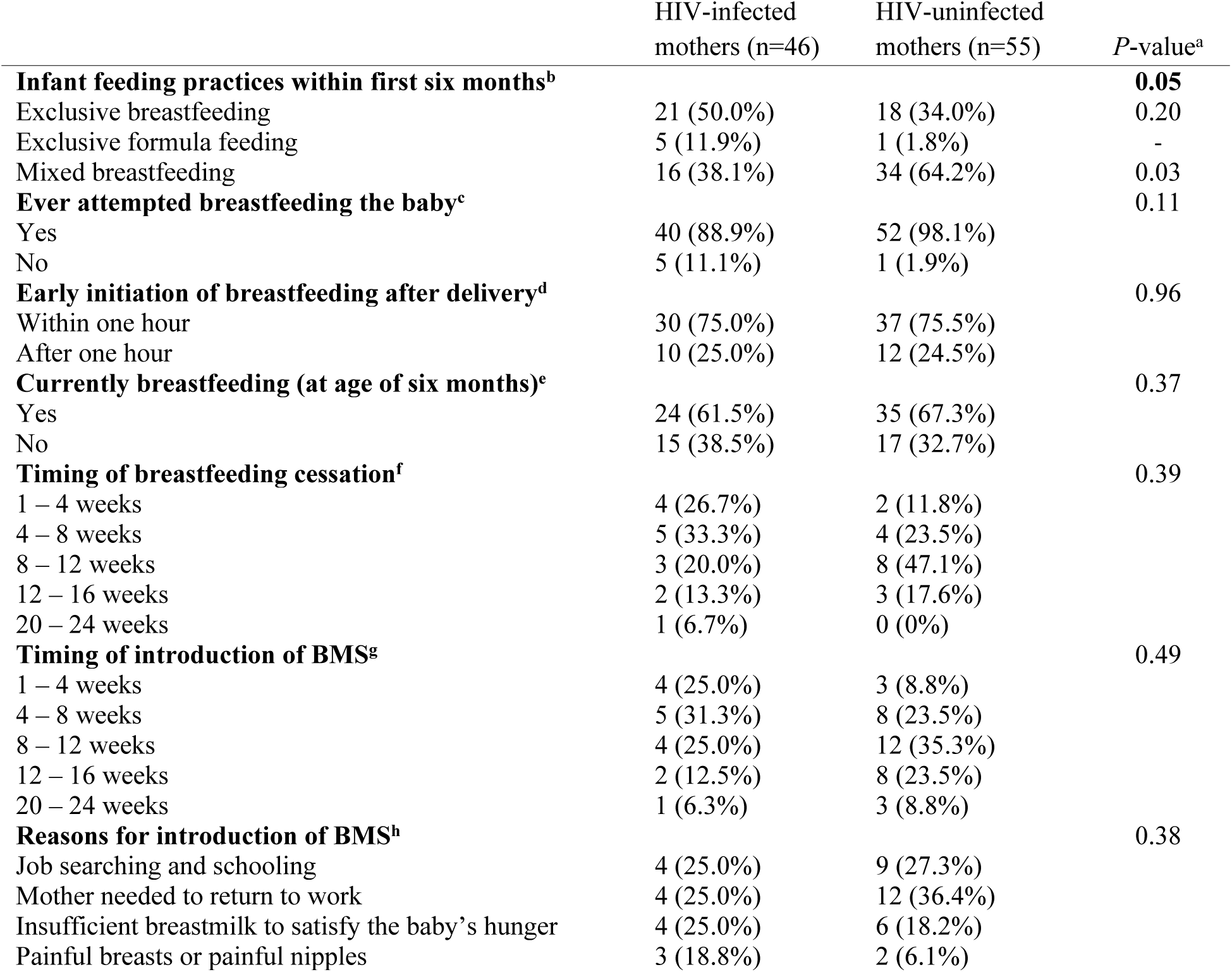
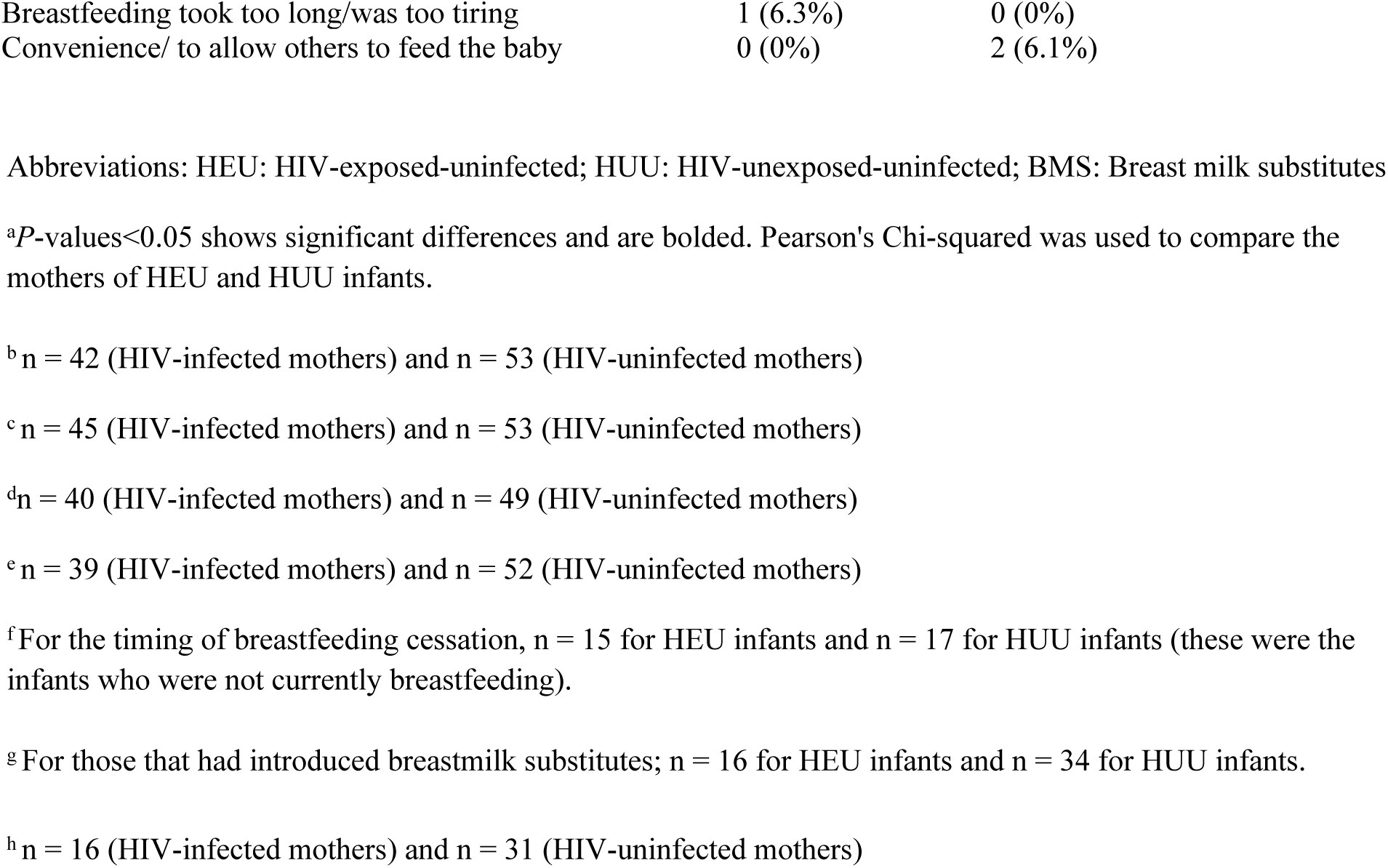
The infant feeding practices of HIV-infected and HIV-uninfected mothers during the first six months of life

|  | HIV-infected<br>mothers (n=46) | HIV-uninfected<br>mothers (n=55) | *P*-value<sup>a</sup> |
| --- | --- | --- | --- |
| **Infant feeding practices within first six months<sup>b</sup>** |  |  | **0.05** |
| Exclusive breastfeeding | 21 (50.0%) | 18 (34.0%) | 0.20 |
| Exclusive formula feeding | 5 (11.9%) | 1 (1.8%) | - |
| Mixed breastfeeding | 16 (38.1%) | 34 (64.2%) | 0.03 |
| **Ever attempted breastfeeding the baby<sup>c</sup>** |  |  | 0.11 |
| Yes | 40 (88.9%) | 52 (98.1%) |  |
| No | 5 (11.1%) | 1 (1.9%) |  |
| **Early initiation of breastfeeding after delivery<sup>d</sup>** |  |  | 0.96 |
| Within one hour | 30 (75.0%) | 37 (75.5%) |  |
| After one hour | 10 (25.0%) | 12 (24.5%) |  |
| **Currently breastfeeding (at age of six months)<sup>e</sup>** |  |  | 0.37 |
| Yes | 24 (61.5%) | 35 (67.3%) |  |
| No | 15 (38.5%) | 17 (32.7%) |  |
| **Timing of breastfeeding cessation<sup>f</sup>** |  |  | 0.39 |
| 1 – 4 weeks | 4 (26.7%) | 2 (11.8%) |  |
| 4 – 8 weeks | 5 (33.3%) | 4 (23.5%) |  |
| 8 – 12 weeks | 3 (20.0%) | 8 (47.1%) |  |
| 12 – 16 weeks | 2 (13.3%) | 3 (17.6%) |  |
| 20 – 24 weeks | 1 (6.7%) | 0 (0%) |  |
| **Timing of introduction of BMS<sup>g</sup>** |  |  | 0.49 |
| 1 – 4 weeks | 4 (25.0%) | 3 (8.8%) |  |
| 4 – 8 weeks | 5 (31.3%) | 8 (23.5%) |  |
| 8 – 12 weeks | 4 (25.0%) | 12 (35.3%) |  |
| 12 – 16 weeks | 2 (12.5%) | 8 (23.5%) |  |
| 20 – 24 weeks | 1 (6.3%) | 3 (8.8%) |  |
| **Reasons for introduction of BMS<sup>h</sup>** |  |  | 0.38 |
| Job searching and schooling | 4 (25.0%) | 9 (27.3%) |  |
| Mother needed to return to work | 4 (25.0%) | 12 (36.4%) |  |
| Insufficient breastmilk to satisfy the baby's hunger | 4 (25.0%) | 6 (18.2%) |  |
| Painful breasts or painful nipples | 3 (18.8%) | 2 (6.1%) |  |

|  |  |  |
| --- | --- | --- |
| Breastfeeding took too long/was too tiring | 1 (6.3%) | 0 (0%) |
| Convenience/ to allow others to feed the baby | 0 (0%) | 2 (6.1%) |

Abbreviations: HEU: HIV-exposed-uninfected; HUU: HIV-unexposed-uninfected; BMS: Breast milk substitutes
<sup>a</sup>*P*-values<0.05 shows significant differences and are bolded. Pearson's Chi-squared was used to compare the mothers of HEU and HUU infants.
<sup>b</sup> *n* = 42 (HIV-infected mothers) and *n* = 53 (HIV-uninfected mothers)
<sup>c</sup> *n* = 45 (HIV-infected mothers) and *n* = 53 (HIV-uninfected mothers)
<sup>d</sup> *n* = 40 (HIV-infected mothers) and *n* = 49 (HIV-uninfected mothers)
<sup>e</sup> *n* = 39 (HIV-infected mothers) and *n* = 52 (HIV-uninfected mothers)
<sup>f</sup> For the timing of breastfeeding cessation, *n* = 15 for HEU infants and *n* = 17 for HUU infants (these were the infants who were not currently breastfeeding).
<sup>g</sup> For those that had introduced breastmilk substitutes; *n* = 16 for HEU infants and *n* = 34 for HUU infants.
<sup>h</sup> *n* = 16 (HIV-infected mothers) and *n* = 31 (HIV-uninfected mothers)

### Common complementary foods introduced to infants

Overall, by six months 89.1% of WLHIV and their HIV-negative counterparts self-reported introducing their infants to BMS and/or complementary foods, with no significant difference between the groups (92.7% of HUU; 84.8% of HEU; *p=*0.20). The most common food items introduced to HEU and HUU infants, respectively, were CIC (48.7% vs 70.6%; *p*=0.03), fruits and vegetables (33.3% vs 15.7%; *p*=0.05) and bottled baby foods (25.6% vs 41.2%; *p*=0.12) (Table 3). The common fruit items were banana and orange, and vegetables included potatoes, sweet potato, and butternut. Margarine, salt, cooking oil, BMS and peanut butter were commonly used during infants’ food preparation for both groups.

**Table 3:**
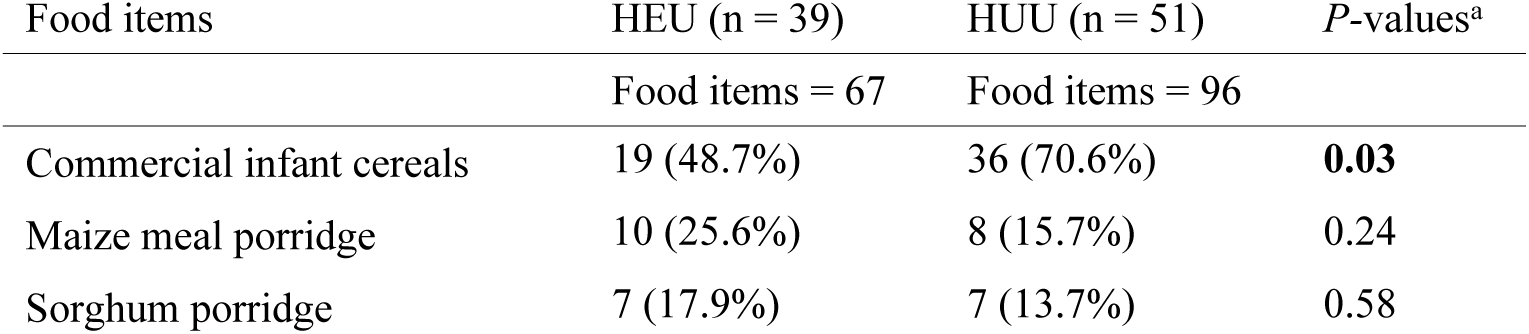
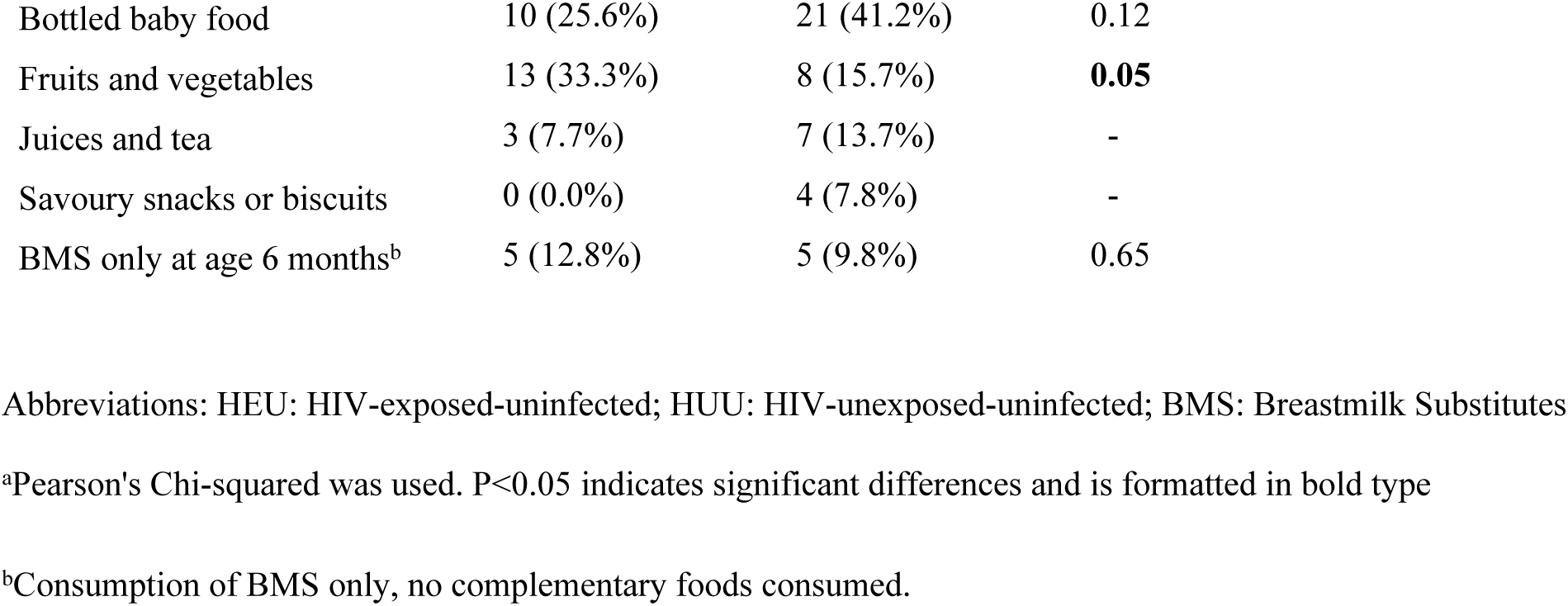
The common complementary foods and breastmilk substitutes given to infants, comparing HIV-exposed-uninfected to HIV-unexposed-uninfected infants

| Food items | HEU (*n* = 39) | HUU (*n* = 51) | *P*-values<sup>a</sup> |
| --- | --- | --- | --- |
|  | Food items = 67 | Food items = 96 |  |
| Commercial infant cereals | 19 (48.7%) | 36 (70.6%) | **0.03** |
| Maize meal porridge | 10 (25.6%) | 8 (15.7%) | 0.24 |
| Sorghum porridge | 7 (17.9%) | 7 (13.7%) | 0.58 |
| Bottled baby food | 10 (25.6%) | 21 (41.2%) | 0.12 |
| Fruits and vegetables | 13 (33.3%) | 8 (15.7%) | **0.05** |
| Juices and tea | 3 (7.7%) | 7 (13.7%) | - |
| Savoury snacks or biscuits | 0 (0.0%) | 4 (7.8%) | - |
| BMS only at age 6 months<sup>b</sup> | 5 (12.8%) | 5 (9.8%) | 0.65 |

Abbreviations: HEU: HIV-exposed-uninfected; HUU: HIV-unexposed-uninfected; BMS: Breastmilk Substitutes
<sup>a</sup>Pearson's Chi-squared was used. $P < 0.05$ indicates significant differences and is formatted in bold type
<sup>b</sup>Consumption of BMS only, no complementary foods consumed.

### The daily cost of food per infant

The calculations considered the total amount of food and BMS consumed as well as the price per 100g for each food item consumed. The total daily mean cost of diet of HEU infants was 8.55±7.35 ZAR ($0.68±0.59 USD) while it was 10.97±7.92 ZAR ($0.88±0.63 USD) for HUU infants, with no significant difference between the groups (*p*=0.10).

### The daily macronutrient intake and their costs per infant

The nutrient intake and cost thereof, as reported in Table 4, indicate no differences in median intakes of protein, fat, and carbohydrate between HEU and HUU infants (*p*-values>0.05), for all complementary foods and BMS consumed in addition to any breastmilk. No differences were found in the cost for these macronutrients between the HEU and HUU groups *(p*-values>0.05). The findings on adequacy of macronutrient intake of complementary diets showed similar low intakes of protein, fat and carbohydrates between HEU and HUU (Table 5). These findings were based on the estimated average requirement (EAR) for protein (1g/kg/d) and adequate intake (AI) values for fat (30g) and carbohydrate (95g) established by the US Institute of Medicine [31].

**Table 4:**
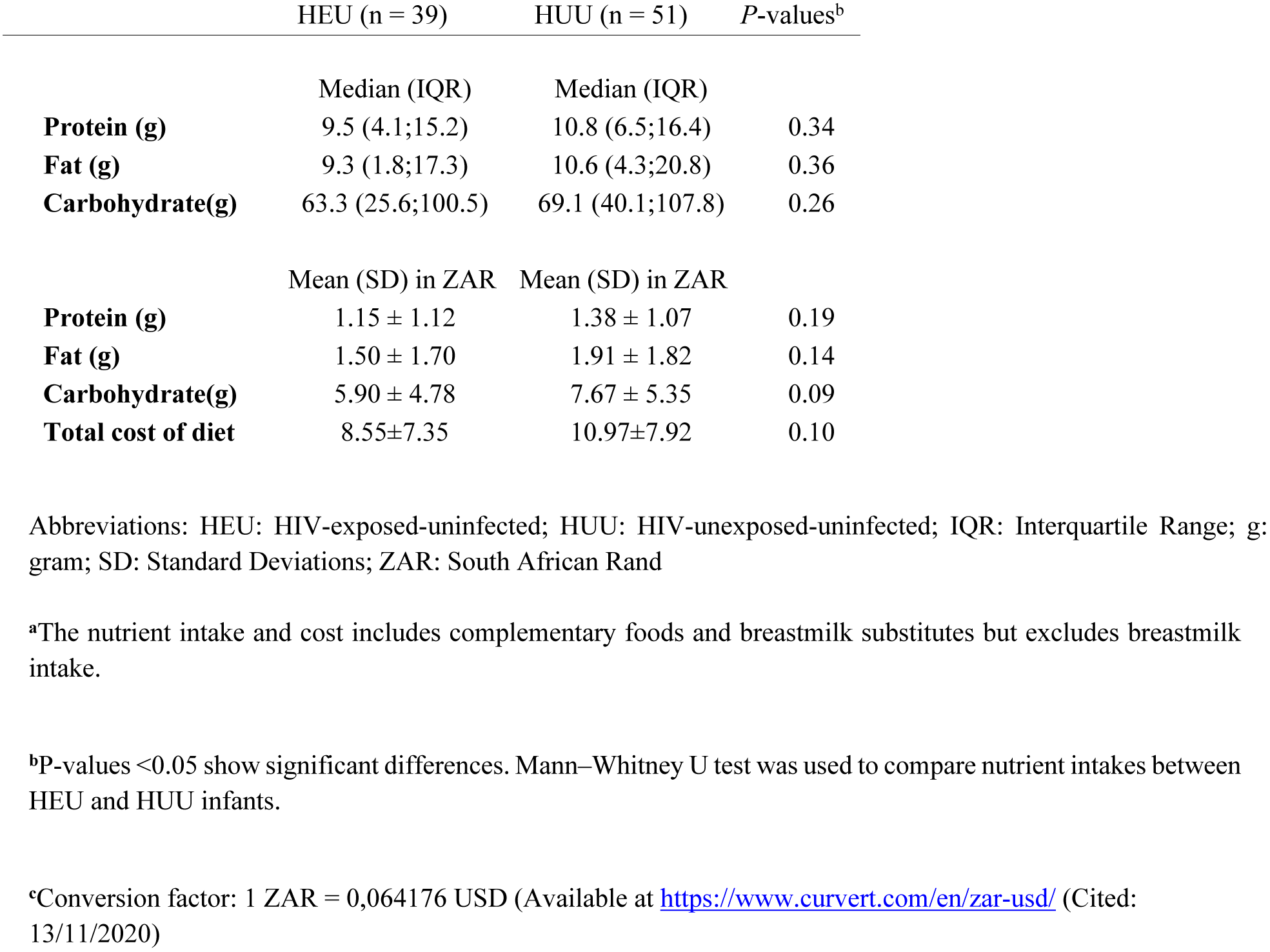
The infants’ nutrient intake (excluding breastmilk intake) and their daily mean cost per day, comparing HIV-exposed-uninfected to HIV-unexposed-uninfected infants^a^

|  | Median (IQR) | Median (IQR) |  |
| --- | --- | --- | --- |
| **Protein (g)** | 9.5 (4.1;15.2) | 10.8 (6.5;16.4) | 0.34 |
| **Fat (g)** | 9.3 (1.8;17.3) | 10.6 (4.3;20.8) | 0.36 |
| **Carbohydrate(g)** | 63.3 (25.6;100.5) | 69.1 (40.1;107.8) | 0.26 |

|  | Mean (SD) in ZAR | Mean (SD) in ZAR |  |
| --- | --- | --- | --- |
| **Protein (g)** | 1.15 ± 1.12 | 1.38 ± 1.07 | 0.19 |
| **Fat (g)** | 1.50 ± 1.70 | 1.91 ± 1.82 | 0.14 |
| **Carbohydrate(g)** | 5.90 ± 4.78 | 7.67 ± 5.35 | 0.09 |
| **Total cost of diet** | 8.55±7.35 | 10.97±7.92 | 0.10 |

Abbreviations: HEU: HIV-exposed-uninfected; HUU: HIV-unexposed-uninfected; IQR: Interquartile Range; g: gram; SD: Standard Deviations; ZAR: South African Rand
<sup>a</sup>The nutrient intake and cost includes complementary foods and breastmilk substitutes but excludes breastmilk intake.
<sup>b</sup>P-values <0.05 show significant differences. Mann–Whitney U test was used to compare nutrient intakes between HEU and HUU infants.
<sup>c</sup>Conversion factor: 1 ZAR = 0,064176 USD (Available at https://www.curvert.com/en/zar-usd/ (Cited: 13/11/2020)

**Table 5:** The daily adequacy of macronutrient intake from complementary diets among HIV-exposed-uninfected and HIV-unexposed-uninfected infants using the dietary reference intake of the Institute of Medicine as comparisons^a^

| Macronutrients | DRI (6-months) | HEU (n = 34) | HUU (n = 46) | P-value<sup>b</sup> |
| --- | --- | --- | --- | --- |
| **Protein<sup>c</sup>** | 1g/kg/day |  |  | 0.89 |
| Above |  | 21.2% | 17.4% |  |
| Below |  | 78.8% | 82.6% |  |
| **Fat<sup>d</sup>** | 30g |  |  | - |
| Above |  | 0.0% | 0.0% |  |
| Below |  | 100.0% | 100.0% |  |
| **Carbohydrate<sup>d</sup>** | 95g |  |  | 0.24 |
| Above |  | 36.4% | 21.7% |  |
| Below |  | 63.6% | 78.3% |  |

Abbreviations: BMS: Breastmilk substitute; HEU: HIV-exposed-uninfected; HUU: HIV-unexposed-uninfected; DRI: Dietary Reference Intake; EAR: Estimated Average Requirement; AI: Adequate Intake; g: gram; kg: kilogram
<sup>a</sup>The macronutrient adequate analysis involved only complementary diets consumed by HEU (N=34) and HUU (n=46) infants.
<sup>b</sup>P-values <0.05 shows significant differences. Pearson's Chi-squared was used to compare nutrient intakes between HEU and HUU infants.
<sup>c</sup>EAR established by Institute of Medicine [31].
<sup>d</sup>AI of the US DRI published by the Food and Nutrition Board for the Health and Medicine Division, formerly Institute of Medicine [31].

## Discussion

The study determined and compared the infant feeding practices and cost of diet in relation to macronutrient intake of six-month-old HEU versus HUU infants. The findings on infant feeding practices within the first six months contribute to the existing literature, with EBF reported at 50.0% among WLHIV; higher than reported in previous South African studies, and lower (34.0%) in HUU infants. The MBF is predominant among HUU infants. The mean total daily diet cost was similar among HEU and HUU infants, as well as the cost and intakes of protein, fat, and carbohydrates.

The WLHIV were found to have lower educational attainment compared to their counterparts, and according to Hargreaves and colleagues, HIV prevalence is higher amongst individuals with lower educational level [32]. Further, maternal HIV infection was associated with lower household monthly income. Naidoo et al. reported a reduced ability to generate revenue in households affected by HIV [33], while Pienaar and co-workers reported a high rate of unemployment among adults living with HIV compared to HIV-negative adults [34]. Lower-income could be linked to lower educational qualifications. We found non-significant differences in terms of food purchasing practices between WLHIV and their HIV-negative counterparts, despite the lower-income status of households of WLHIV. However, the overall amount of food purchased was not studied, so it may be that due to food insecurity households of WLHIV bought less food. The economic status of a household is the key determining factor for household food security as it directly impacts the food purchasing power and practices [18]. A study that determined household food security reported high levels of moderate and severe food insecurity among households affected by HIV infection [35]. The difference with the current study may be due to the small household sizes of WLHIV, as most had few children.

In our study, the self-reported percentage of early initiation of breastfeeding (EIBF) was high in both WLHIV and HIV-negative women (75.0%; 75.5%), and higher than previously reported in WLHIV (56.6%), HIV-negative women (61.2%) and the general population (67.3%) in South Africa [15,36], as well as globally in the general population (57.6%) as stated in a WHO global survey [37] and 58.3% in Ethiopia [38]. The EIBF rates also surpassed the WHO global target of 70% in both groups. The percentage of ever attempted breastfeeding was also higher in both groups compared to the previously reported 57.1% and 80.9% in South Africa [36,39]. On the other hand, EBF was on average (50.0%) in WLHIV; the 50% target of the United Nations Decade of Nutrition, but lower (34.0%) than this target in HIV-negative women. The reported percentage of EBF in this study among WLHIV is high compared to previous South African studies. These include the reported 12% in 2013 and 13% in 2015 the general population [40,41], and 35.6% as well as 44.7% reported among WLHIV and 42.8% in HIV-negative women [42,43]. The findings differed from those reported in separate studies among Ethiopian WLHIV (63.4% and 83.8%) [44,45]. The EBF and MBF practices may be influenced by the availability of a supportive society and the impact of aggressive marketing by the BMS industry on breastfeeding practices [46]. The percentages of MBF reported in this study was comparatively higher than previously reported in South Africa (12.4%) and from Ethiopian (10.5%) [42,45]. Mothers may assume that breastmilk is not sufficient for the infants and mixed feed to supplement it, and this places infants at risk of vertical HIV transmission as their intestinal mucosa is still immature and not well equipped to handle additional foods [45]. The FF reported in this study was lower than the previously reported percentages in South African WLHIV with infants aged 3 to 6 months [15,42]. Attending support group on infant feeding practices and proper knowledge on vertical transmission of HIV through breastfeeding is associated with FF [42].

Most (89.1%) infants in the study were introduced to complementary feeding and BMS, while a few (10.9%) were still exclusively breastfed. The Ethiopian study reported 60.5% and 19.0% of timely and early introduction of complementary feeding, respectively [47]. A higher proportion of HUU infants were consuming CIC compared to their HEU counterparts. High consumption of CIC has been documented in South Africa among six months old infants; 70% by Swanepoel et al. and 92% by Budree et al. The possible reason for lower intake among HEU infants may be due to the cost of these food items, as consumption hinges on affordability [48]. Intakes of staple food items such as maize meal and sorghum porridge did not differ between the HEU and HUU groups. These findings are similar to previously reported 23% and 20% in South Africa [20,41]. Maize meal and sorghum porridge are well known common traditional weaning foods in sub-Saharan Africa [49], are readily available at low cost. Although the fortification of maize meal is mandatory in South Africa, the consumption of maize meal porridge is still of concern among children as they consume a small quantity that is highly diluted with water, resulting in poor quality [19].

The findings showed non-significant differences between the HEU and HUU infants in terms of the total daily mean cost of diet, albeit that we documented a lower cost of diet in the HEU infants compared to the HUU infants (8.55±7.35 ZAR ($0.68±0.59 USD) vs 10.97±7.92 ZAR ($0.88±0.63 USD)), considering the lower intake of CIC by HEU infants than their counterparts. Assumption is that the included BMS may be accountable for the diet cost discrepancy as formula milk may be costly. A daily Kenyan diet for 6 – 8 months old infants is estimated to cost a total of 17.04 ZAR ($1.363 USD), this includes the cost of combined energy only, micronutrient nutritious and food habits nutritious diets, utilising the Cost of the Diet software [50].

We found non-significant differences between the HEU and HUU infants for the cost of macronutrient intakes. There were no reports in the literature on the cost of nutrients in a complementary diet for the six-month age group. The percentages of adequate intake of protein and carbohydrates were low in both groups, and fat intake had the lowest percentage. However, 61.5% of HEU and 67.9% of HUU infants were still being breastfed at the time of the study visit, with the breastmilk providing additional macronutrients. It is possible that infants were recently introduced to complementary foods and were consuming more of breastmilk and BMS than foods. Most mothers did not adhere to the EBF recommendation, feasibly introducing complementary feeds early, with financial implications in addition to being nutritionally suboptimal. The findings for protein and carbohydrate differed from the Malawian study, which reported adequate intakes among non-breastfed HEU infants [51]. However, the finding for low fat intake was in line with the Malawian study [51]. The inadequate macronutrient intakes in this study may be because the infants were recently introduced to complementary feeding.

The study determined the estimated daily cost of the complementary diet and BMS of HEU infants, who are likely to be vulnerable to food insecurities and are reported to be at greater risk of poor development [2]. The researcher obtained information or responses from caregivers about complementary foods and ways of enriching family foods given to infants through the infant’s 24-hour recall. We determined the daily macronutrient intake of complementary diet and BMS as they contribute to the cost of diet. The BMS was included as it contributes to the cost of diet to the family, but this had limited the exactness of the estimation of the cost of complementary diets. For future research, measuring the nutrient intakes and their cost should incorporate the anthropometric measurements, so to determine if there is any association between the feeding practices and the infants’ nutritional statuses.

## Conclusions

We found no significant differences in cost of feeding and macronutrient intakes between HEU and HUU. Suboptimal breastfeeding practices remains an issue within first six months with no differences in EBF between HEU and HUU children and MBF being dominant in HUU children. Suboptimal EBF are likely linked to maternal misconceptions about infant feeding practices such as breastmilk alone being not enough for the infants or HIV vertical transmission. More sustained effort is required to support and promote EBF among South African mothers.

## Data Availability

Data is available upon request

## Acknowledgements

This is the substudy to Siyakhula study. We thank our research assistants: Maryjane Ntima, Sheilla Sono and Kedibone Matshai; Phumudzo Mamphwe for assistance with data cleaning; and the mothers and their children for their participation.

